# Meningococcal epidemiology in Ethiopia after MenAfriVac: A situational analysis of surveillance gaps and national meningococcal vaccine policy

**DOI:** 10.64898/2026.09.09.26362624

**Authors:** Lemma Demissie Regassa, Caroline Trotter, Nega Assefa, J. Anthony G. Scott

**Affiliations:** London School of Hygiene and Tropical Medicine, London, UK; Haramaya University, Dire Dawa, Ethiopia; Hararghe Health Research (HHR), Haramaya University, College of Health and Medical Sciences, Harar, Ethiopia; University of Cambridge, Cambridge, UK; Imperial College London, London, UK

**Keywords:** Meningitis epidemics, Meningitis belt, Ethiopia, Serogroup A meningococcal disease, MenAfriVac vaccine, Vaccine protection waning

## Abstract

Ethiopia introduced the MenAfriVac vaccine to control epidemics of serogroup A meningococcal disease (MenA) between 2013 and 2015. Since the campaign, no MenA epidemics have been reported. However, lacking a follow-up strategy, two critical questions remained unanswered: (1) is protection against MenA waning to a point where epidemics could re-emerge and (2) are non-A serogroups, known to cause outbreaks elsewhere in the meningitis belt, currently circulating in Ethiopia? To inform future policy, this situational analysis draws on historical records, post-2015 surveillance data, MenAfriVac campaign reports, and the current policy landscape to assess meningococcal epidemiology in Ethiopia.

Available national data cannot answer either question. Disease notification relies on sentinel surveillance at three hospitals; none located in the epidemic risk western regions and none with a defined catchment population. Cases are rarely laboratory-confirmed, almost never serogrouped, and the resulting data are not reported in a timely manner. Primary carriage and serosurvey studies remain limited, and their methodological heterogeneity including diverse populations, sampling techniques and laboratory assays restricts their generalisability for population-level policy decisions. In the absence of quality primary data, indirect risk estimation based on campaign coverage and regional waning models indicates that population protection dropped from 85% in 2015 to 24% in 2024 and will fall further to 11% by 2030 without intervention. Despite this, the current E-NITAG prioritisation exercise relies on this sparse evidence base, excludes MenAfriVac from the candidate list, and does not incorporate costing or serogroup-coverage data.

Addressing the meningococcal evidence gap requires rebuilding national surveillance into a tiered system. Under this structure, primary facilities manage clinical case detection, regional and zonal hospitals perform confirmatory testing, and representative tertiary hospitals conduct serogrouping and genomic characterization with national laboratory support. Until this longitudinal surveillance yields sufficient data for policy decisions, supplementary carriage and seroepidemiological studies remain necessary to assess circulating serogroups and population immunity.

## Introduction

Ethiopia is on the eastern edge of the African meningitis belt. Since 1901, the country has experienced large meningococcal epidemics every 4 to 12 years (1), mainly caused by meningococcal serogroup A (MenA). To interrupt these recurrent epidemics, Ethiopia conducted mass vaccination campaigns with MenAfriVac targeting 1-29 year olds from 2013 to 2015(2). Mirroring successes across the African meningitis belt, this intervention successfully eliminated major MenA outbreaks in Ethiopia; no MenA epidemic has been recorded since 2015 (3).

Although the MenAfriVac campaign was reported to have achieved remarkably high coverage, protection is expected to decline over time (4, 5). The protected proportion of the population declines through two mechanisms simultaneously: antibody waning in vaccinated individuals and the accumulation of unvaccinated birth-cohorts. Modelling from other meningitis belt countries indicates that, without follow-up vaccination, populations could return to a highly vulnerable state within a decade (6, 7). Ethiopia has not used MenAfriVac in campaigns or routine immunisation since 2015 and may therefore be vulnerable to a resurgence of MenA outbreaks.

In addition to the risk of resurgent MenA outbreaks, surveillance and outbreak reports since 2015 have described localised increases in disease attributed to serogroups W, X and C (8, 9). Elsewhere in the belt MenC and MenW have caused large epidemics in the post-MenAfriVac period (10–13). This shift in outbreak-causing serogroups exposes the country to a dual risk: MenA epidemics could re-emerge while other serogroups could continue to circulate unchecked.

At the same time, the existing serological and carriage evidence is fragmented and geographically limited, so Ethiopia lacks the nationwide data on circulating serogroups, population immunity and long-term vaccine impact needed to choose between preventive strategies. In Ethiopia, the Ministry of Health (MoH) relies on evidence-informed prioritisation through the Ethiopian National Immunization Technical Advisory Group (E-NITAG) and the national immunisation strategy; however, this evidence gap has prevented clear prioritisation of meningococcal vaccines. A situational analysis that integrates historical epidemiology, contemporary trends, immunity gaps and policy status may therefore inform national decision-making and guide future meningococcal control strategy.

## Methods

This situational analysis employed a mixed-methods approach comprising a desk review, secondary quantitative data analysis, modelling of population immunity and a policy environment assessment. The study was approved by Haramaya University, National Ethics Review Board (Ethiopia) and London School of Hygiene and Tropical Medicine (LSHTM).

### Data sources

Quantitative data were triangulated from three sources. Epidemic data before 2012 were obtained from guidelines and reports held by the MoH and Ethiopian Public Health Institute (EPHI)(14). Disease notifications were obtained from the WHO meningitis dashboard (15). Sentinel disease surveillance data from Tikur Anbessa Specialized Hospital (TASH), Yekatit 12 Hospital (Y12H) and University of Gondar Comprehensive Specialized Hospital (UoGH), collected between 2012 and 2020, were obtained from EPHI. Both TASH and Y12H are in Addis Ababa, the capital city (16, 17). UoGH, a referral hospital in Gondar town, is located in northwest Ethiopia (18, 19). Data for the MenAfriVac mass vaccine campaign were obtained from the MoH. These data were aggregated at district level and report the doses administered and the demographic profile of the vaccinated population. An adjusted population denominator from the Ethiopian Statistical Services was used to estimate MenAfriVac coverage.

The policy-environment analysis drew on publicly available sources: E-NITAG’s prioritisation document (20), WHO’s Strategic Advisory Group of Experts on Immunization (SAGE) recommendations (4, 21), and Gavi funding guideline (22). To clarify decision sequences and identify relevant materials, exploratory informal discussions were held with national Expanded Programme on Immunization (EPI) head and E-NITAG members. These discussions were purely contextual, no interview schedules were used, no personal data were recorded, and no individuals are quoted or identified.

Published and online evidence was sought from databases (PubMed, Embase, and Web of Science), grey literature, Ethiopian journals and university repositories. We used a combination of Medical Subject Headings (MeSH) and free-text terms including (meningococcal OR *Neisseria meningitidis* or *N. meningitidis*) AND (carriage prevalence OR seroprevalence OR serogroup shift OR MenAfriVac OR vaccine impact) AND (Ethiopia OR African meningitis belt).

### Operational definitions

To avoid ambiguity, the following terms are used consistently throughout this analysis. These terms are defined based on National Meningococcal surveillance and management guideline definitions (14) and updated WHO guideline (23):

- Disease notification: a suspected meningitis case report entering the routine surveillance system.
- Suspected case: Sudden fever (>38.5 °C rectal / 38.0 °C axillary) with neck stiffness, altered consciousness, or meningeal signs (bulging fontanel, convulsions).
- Probable case: Suspected case with turbid/purulent cerebrospinal fluid (CSF) or Gram-negative diplococci on microscopy.
- Confirmed case: Suspected or probable case with *N. meningitidis* identified in CSF/blood via culture, PCR, or latex agglutination (including serogroup if identified).
- Alert Threshold: Triggered by five cases in a single week, or a doubling of cases over three consecutive weeks.
- Epidemic Threshold: Defined as a weekly attack rate exceeding ten suspected cases per 100,000 population per week at the district level for consecutive weeks.
- Zone: The second-largest administrative subdivision in Ethiopia and in this document used to spatially map the protection. Zones are subdivisions of regions and are further divided into woredas (referred to in this document as districts).

### Quantitative analysis

Evidence from the literature review was synthesised to provide historical context.

Vaccine coverage and population protection were estimated using MenAfriVac uptake reports. The target population comprised individuals aged 1–29 years eligible for vaccination during the campaigns. Coverage was calculated by dividing total doses administered by the target population. Annual population denominators (ages 1–29) (*N_t_*) were derived from Ethiopian Statistical Service (ESS) projections based on the 2007 census (24). The vaccination campaign was launched first in the high-risk regions in 2013, followed by the medium-risk in 2014, and the low-risk in 2015. The campaign targeted 18,926,853 eligible individuals in Phase 1; 26,910,795 in Phase 2; and 16,190,737 in Phase 3.

The protected proportion declines through two independent mechanisms: waning of vaccine-induced immunity within the 2013–2015 cohorts and dilution of protected cohorts as unvaccinated individuals enter (*N*_*t*_). Based on evidence from Yaro et al. (7) and White et al.(6) indicating age-specific duration, waning of MenAfriVac protection was modelled assuming an exponential decay with an average duration of 5 years (20% waning rate) for children under 5 and 10 years (10% waning rate) for individuals aged 5 and older. Demographic shifts were incorporated through two aging dynamics: entry into the target by surviving infants (reached 1 year) projected by the ESS and exit upon reaching age 30. The annual exit rate was calculated by dividing the number of individuals turning thirty by the total target population (1–29 years) during the same period. The net protected population (P) was defined as individuals aged 1-29 years who have been vaccinated with MenAfriVac in whom protection has not waned.

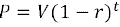

Where P is the protected population at a given time (t), V is the initial protected population following vaccination, and r is the age specific waning rate based on a given duration of vaccine protection (i.e., r=1/average duration of protection). The model was further adjusted using the cohort survival (1-exit ratio).

The impact of assumptions regarding the duration of vaccine protection was investigated on the core age groups as classified during the MenAfriVac post-mass campaign survey, including 1-29 years, 5-15 years, 16-29 years and the under 5-year-olds (highest burden age group). We also compared the simulated protected population and projected population size aged 1 to 29 years until 2030 to indicate the gap between protected population and high-risk (1-29 years old) populations (either for disease or carriage). We also simulated the proportion of protected population per district, assuming the age-specific waning rate and mapped at the zonal administrative level. To account for the uncertainty around the duration of protection, we also considered the average duration of 10 years, and 20 years in sensitivity analyses using modelling study approach (25).

Individuals born after the campaign were assumed to be unvaccinated and susceptible. Given evidence of robust herd protection and minimal post-2013 transmission across the meningitis belt (26, 27), we excluded natural immunity from carriage or recovery. The model also omitted vaccinated migrants and assumed net migration had no significant impact on age structure. All calculations and simulations were conducted using R version 4.4.1 and Microsoft Excel. The geographical difference map was created with ArcGIS Pro 3.1.

### Policy-environment analysis

We evaluated several policy frameworks, including Walt and Gilson’s health-policy triangle (28), Kingdon’s multiple-streams model (29), Shiffman and Smith’s political-priority framework (30), Sabatier’s advocacy-coalition framework (31) and the linear policy-cycle heuristic (32). Although these frameworks are robust for agenda-setting and advocacy analysis, they require richer process data than were available. Consequently, the health-policy triangle was selected for its alignment with Ethiopia’s multi-stakeholder landscape and its capacity to leverage scarce data. The framework examines four dimensions: actors (the Ethiopian MoH, EPHI, and E-NITAG as primary actors, alongside WHO, Gavi, and Program for Appropriate Technology in Health (PATH) as secondary technical/financial partners); content (national immunisation strategies, epidemic-prevention plans, policy documents); context (historical and environmental disease drivers); and process (decision-making and prioritisation mechanisms by E-NITAG) (Figure 1).

**Figure 1:**
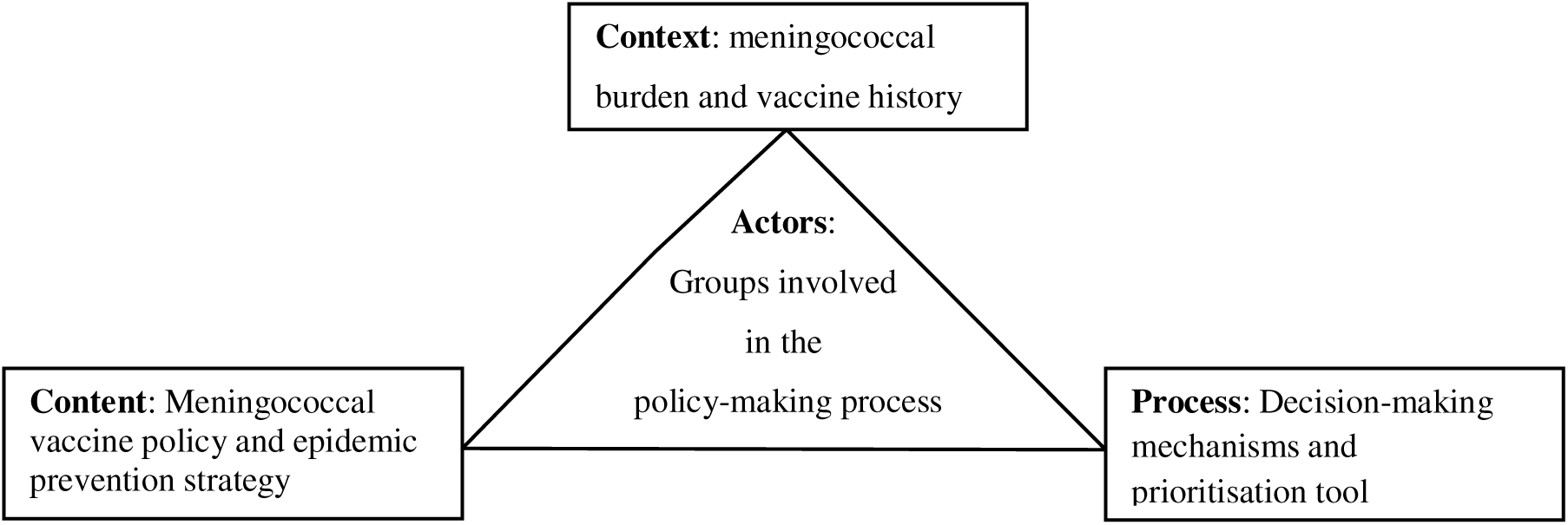
The Walt and Gilson health policy triangle applied to meningococcal vaccine policy in Ethiopia.

## Results

### The Disease epidemiology

#### i. Pre-MenAfriVac era

The first meningococcal epidemic in Ethiopia was documented in 1901 (33). Since then, the pattern of epidemics has mirrored that seen throughout the meningitis belt, typically peaking during the dry season (January–May) and driven primarily by MenA (14, 34). The geographical distribution of the disease has shifted over time. Prior to 1980 outbreaks were reported from the western and northwestern regions; after 1981, the epidemic region expanded south and eastward to include central and southern areas (14, 35). Over 50% of historical cases occurred in children aged <14 years, with just over 40% in persons aged 15 44 years (35–37). Lowland pastoralist regions remained unaffected by large-scale outbreaks (Table 1).

**Table 1:** Meningococcal meningitis epidemiology in Ethiopia from 1900 to 2012.

| Period/Year | Estimated Severity | Key Events / Observations | Caused by | Source |
| --- | --- | --- | --- | --- |
| 1900–1930s | Intermittent | Linked to drought and regional instability (political transition) | MenA | (1, 33, 36–38) |
| 1935–1945 | High burden | Large outbreaks during WWII; surveillance was limited. |  |  |
| 1950s | Moderate | Urban centres saw the first consistent clinical documentation. |  |  |
| 1964 | Epidemic | Highland regions (northwest) hit hard; estimated attack rates among the highest in the belt. |  |  |
| 1981 | Epidemic | Over 50,000 cases estimated. |  |  |
| 1988–1989 | Epidemic | Massive outbreak (~45,000–50,000 cases) |  |  |
| 1996 | Epidemic | Part of the "Great African Wave"; thousands of deaths across rural Ethiopia. |  |  |
| 2000–2003 | Epidemic | MenA and MenC detected; outbreaks hit Addis Ababa and South Regions. | MenA<br>MenC |  |
| 2003–2006 | Localized Surge | 2003–4: Recorded a total of 3326 cases and 160 deaths, 2005: a total of 1061 cases with 46 deaths were reported from four regions, 2006: a total of 3000 cases from all Regions and 43 deaths were reported from three regions, namely Oromia, South regions and Tigray | MenA | (8, 39–41) |
| 2007–2012 | Endemic | Median of 1,056 cases annually<br>Preparation for the new conjugate vaccine began.<br>Baseline surveillance improved as Ethiopia joined the WHO Global Invasive Bacterial Vaccine-Preventable Diseases (IB-VPD) Surveillance Network. |  |  |

Based on the disease burden and history of outbreaks, MoH classified the regions into three broad areas of risk (43). The western and northwestern regions were at high risk, central and south at medium risk and the eastern and lowland regions at low risk. Based on this risk classification, a mass vaccination campaign was implemented using three phases in these three regions from 2013 to 2015 (**Error! Reference source not found**.).

**Figure 2:**
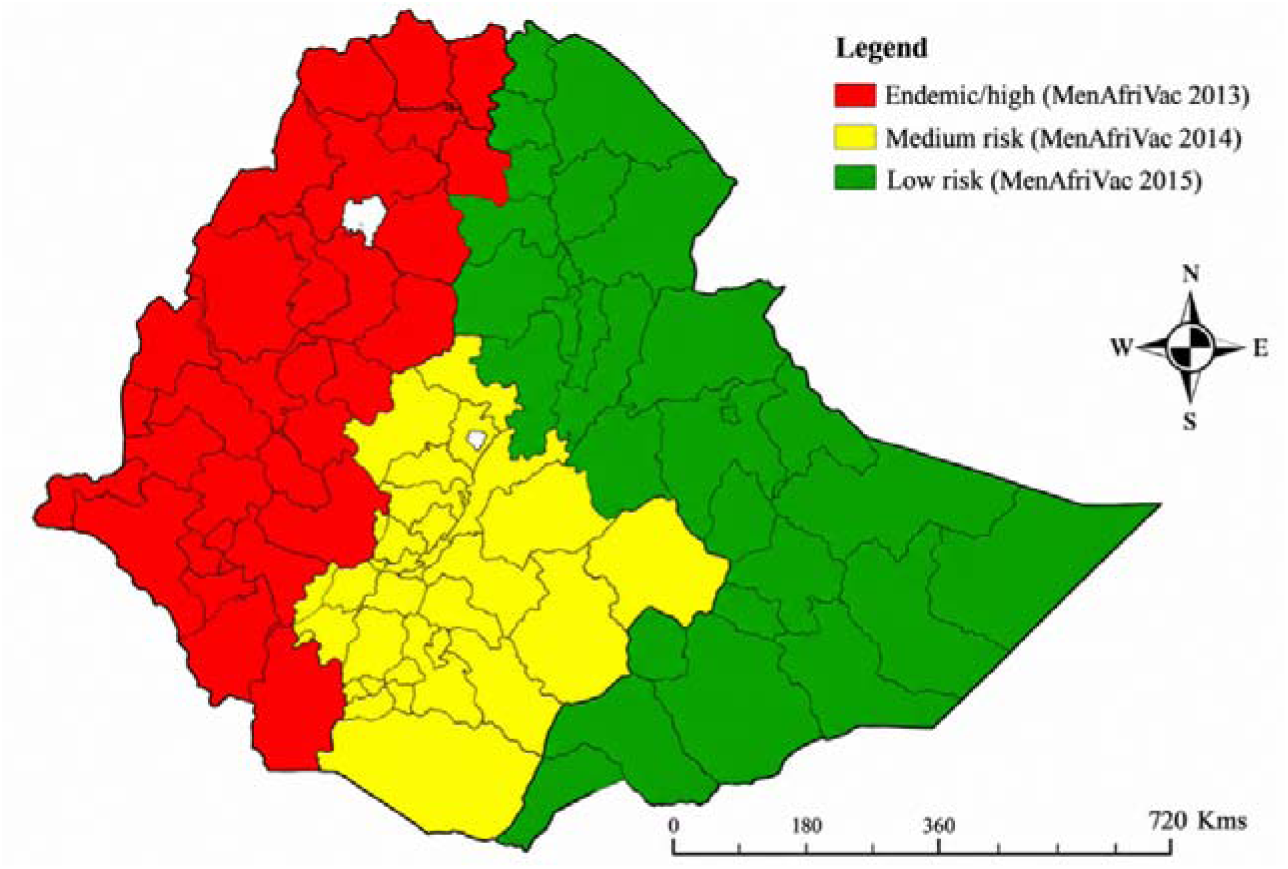
Meningitis risk classification of Ethiopia updated in 2013 (source: Technical Report of MenAfriVac mass vaccination Campaign from 2013 to 2015 (43); The shape file is downloaded from GADM (44))

#### ii. Post-vaccination epidemiology

Following the 2013–2015 campaigns across high, medium, and low-risk regions, no meningococcal outbreaks have crossed the epidemic threshold. However, this impact cannot be definitively verified due to the absence of population-representative laboratory confirmation and serogroup-specific surveillance. Existing data are fragmentary, low-volume, and inadequate to characterise the epidemiology or circulating serogroups of meningococcal disease.

Two data sources reporting meningitis in Ethiopia were identified: the national disease notification system, which reports to WHO, and sentinel surveillance. The notification system compiles clinically reported cases weekly and includes results of CSF testing(14), although most laboratory confirmations originate from the sentinel sites. The two systems are discordant: the notification system reports fewer cases than the pooled totals from the three sentinel hospitals.

The dashboard (15) shows that reporting from Ethiopia has been inconsistent and frequently late. Ethiopia sits at the very bottom of the belt in surveillance volume, with only 90 CSF samples collected and tested since 2016, compared with tens of thousands in other belt countries. Over the same period, the highest post-MenAfriVac meningitis case count was recorded in 2021, with 3,153 suspected cases and 24 deaths, while in 2022 only 5 laboratory-confirmed bacterial meningitis cases were meningococcal.

Together these figures reflect both the small number of sentinel sites submitting samples and the low frequency of laboratory testing (Figure 3).

**Figure 3:**
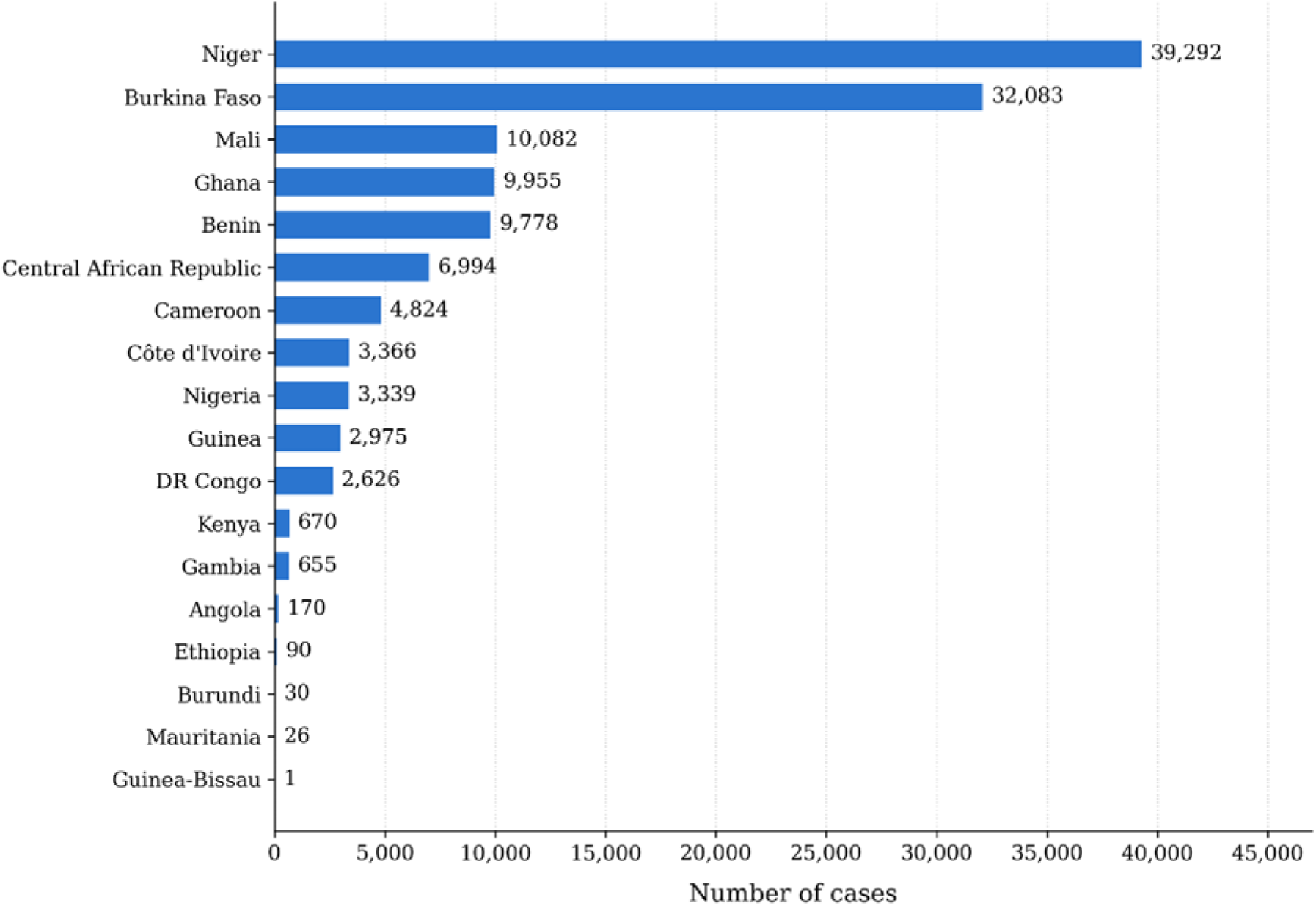
CSF samples tested by country, meningitis belt (data source: WHO Meningitis Dashboard (15)).

Between 2020 and 2026, national weekly epidemiological reports recorded peak clinical meningitis cases during the dry season (weeks 11–19; March–May), alongside an isolated peak in week 29 of 2021. However, the observed seasonal variation was weaker than in typical meningitis belt epidemics. This reduced signal reflects reporting location bias, as sentinel sites are predominantly situated in urban central and northwestern settings rather than the epidemic prone regions, where climatic conditions drive seasonal transmission (Figure 4).

**Figure 4:**
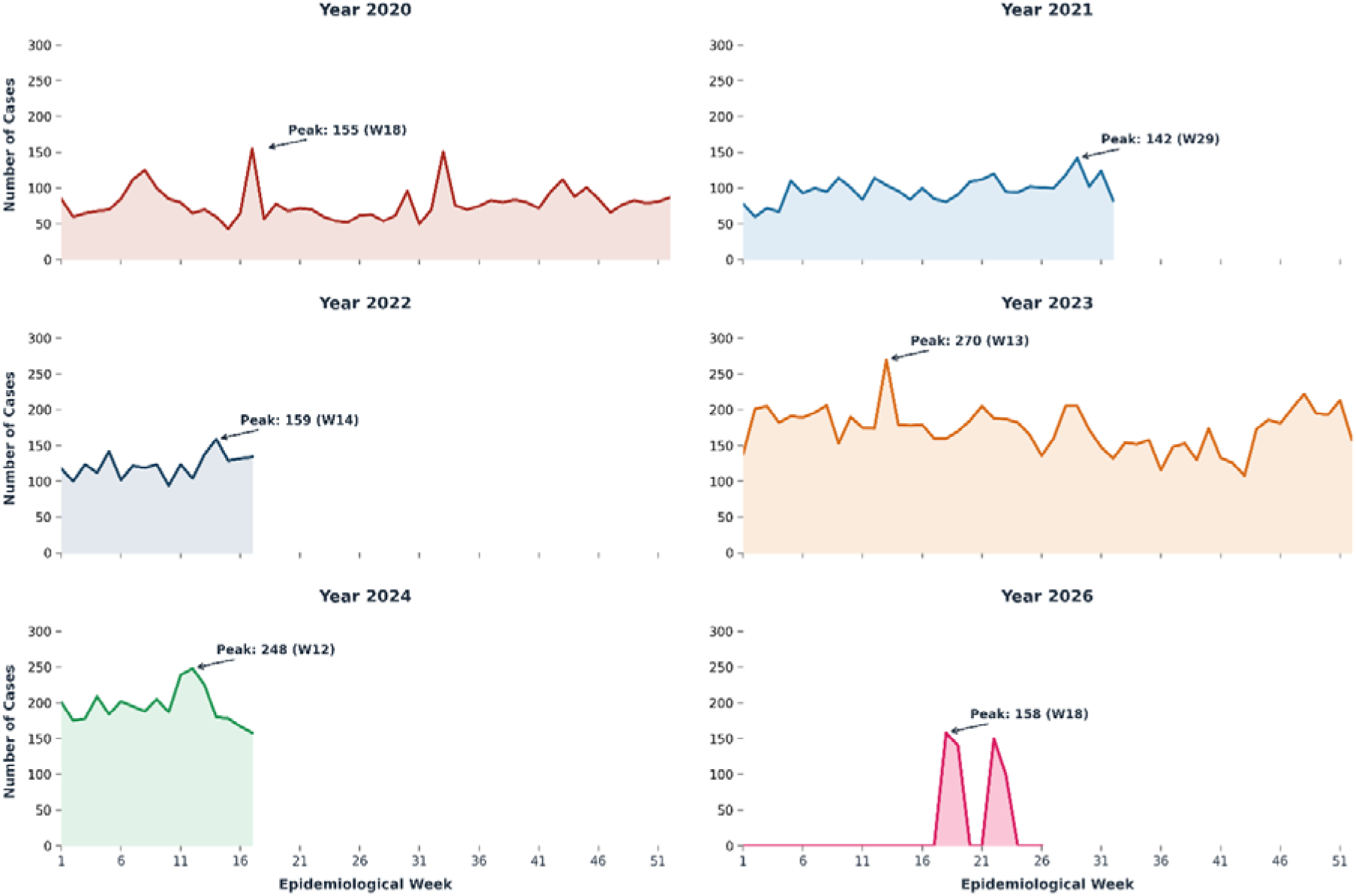
Meningitis cases by WHO’s meningitis week according to the national report to WHO.

Sentinel surveillance, by contrast to the dashboard totals, cultured 5,195 CSF samples in 2012; annual volume subsequently declined to 845–1,168 samples between 2013 and 2019 and dropped to 433 in 2020. Since 2013, confirmed bacterial meningitis cases ranged from 6 to 36 annually, representing confirmation yields of 1%–4%. Among all *N. meningitidis* identified since 2012, only 37 isolates underwent serogrouping and there were no meningococcal cases identified in 2019 or 2020 (Table 2).

**Table 2:** Bacterial meningitis confirmations from sentinel surveillance, Ethiopia, 2012–2020.

| Year | Total tested | *H. influenzae* | *S. pneumoniae* | *N. meningitidis* | Other organisms | Total confirmed meningitis | Proportion of *N. meningitidis* |
| --- | --- | --- | --- | --- | --- | --- | --- |
| 2012 | 5195 | 63 | 69 | 67 | 20 | 219 | 30.6% |
| 2013 | 1085 | 3 | 3 | 0 | 5 | 11 | 0.0% |
| 2014 | 931 | 1 | 6 | 1 | 9 | 17 | 5.9% |
| 2015 | 845 | 4 | 9 | 5 | 18 | 36 | 13.9% |
| 2016 | 942 | 1 | 8 | 3 | 18 | 30 | 10.0% |
| 2017 | 881 | 2 | 2 | 2 | 19 | 25 | 8.0% |
| 2018 | 1168 | 0 | 5 | 2 | 13 | 20 | 10.0% |
| 2019 | 1165 | 0 | 3 | 0 | 15 | 18 | 0.0% |
| 2020 | 433 | 0 | 0 | 0 | 6 | 6 | 0.0% |

National pooled serogrouped data were sparse across the observation period: only 37 serogrouped isolates were reported since 2012 with no data recorded after 2017. MenW (n=20) and MenA (n=12) accounted for most of the isolates recovered. Given the small sample size and the reporting gaps, these counts reflect opportunistic serogroup detection rather than the population-level distribution or dominance of serogroups. Because the current testing volume is too low to detect low-level circulation, the absence of MenA since 2017 does not prove the serogroup has been eliminated (Figure 5).

**Figure 5:**
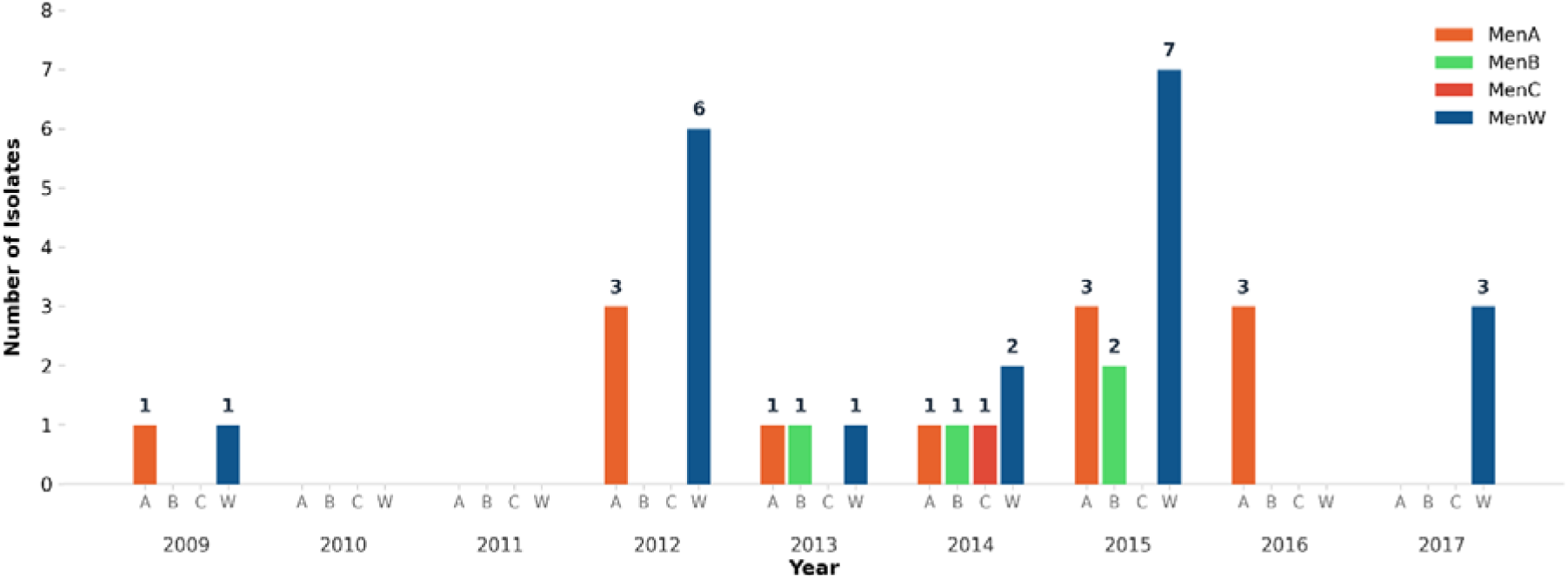
Annual number of isolates of meningococcal meningitis by serogroup pooled from disease notification and sentinel surveillance.

Whereas the disease data of earlier decades, though imperfect, were broadly consistent with belt-wide epidemic patterns and gave a usable picture of disease, the data generated since 2013 fail to provide a reliable epidemiological picture of meningococcal disease in Ethiopia. This inadequacy stems directly from major surveillance limitations. Specifically, laboratory testing is confined to three urban hospitals outside high-risk western regions, systems report conflicting case totals, and peak post-campaign clinical reports lack aetiological confirmation. Furthermore, low culture yields obscure the true disease burden, indicating that years with zero reported detections reflect surveillance breakdown rather than pathogen clearance. Infrequent and inconsistent testing, indicated by a total absence of serogrouped reports since 2017, further prevents the detection of strain replacement or persistent MenA transmission. Clinical surveillance cannot determine whether MenA continues to circulate or population immunity persists. Consequently, evaluating indirect evidence in this setting is essential. This approach combines carriage data to monitor ongoing transmission with serology and modelling based on vaccine uptake to measure remaining protection.

### Nasopharyngeal carriage

As of September 2026, seven meningococcal carriage studies in Ethiopia report wide-ranging prevalence from 6% in the general population (45, 46) to 20% in high-risk groups (47–49). This variability largely stems from population differences; high-risk cohort settings involve closed or semi-closed environments where crowding facilitates transmission. Methodological variations also influenced detection sensitivity, specifically sampling site (nasopharyngeal vs. oropharyngeal) and diagnostic approach (culture alone vs. combined culture and molecular assays). Furthermore, molecularly confirmed pre-MenAfriVac studies (45, 46) and a survey conducted six years post-campaign (50) detected no serogroup A carriage. In contrast, surveys conducted after the mass vaccination campaigns in closed or semi-closed settings (47–49) reported MenA. However, these studies relied exclusively on seroagglutination methods, which are known for poor serogrouping accuracy (51) (Table 3).

**Table 3:** Carriage studies conducted in Ethiopia from 2010 to 2026.

| Author & Year | Study & Population | Sampling Period | Specimen Type | Carriage Prevalence (95% CI) | Serogroups Detected |
| --- | --- | --- | --- | --- | --- |
| MenAfriCar Consortium (2015)(45) | MenAfriCar Baseline, central Ethiopia (Butajira, 3 surveys in dry, rainy and mixed season, age:1-29 yrs; n=5,970) | 2010–2012 | OP | 5.7% (4.6–7.0); 6.1% (4.9–7.5); 7.2% (5.8–8.8) across three surveys | NG dominant: W, X, Y, B, C detected; A 0% |
| Bårnes *et al.* (2016) (46) | Southern Ethiopia Pre-MenAfriVac (Arba Minch, ages:1-29yrs, n=7,479) | 2014 | OP | *N. meningitidis*: 6.6% (6.0-7.2%)<br>*N. lactamica*: 28.1% (27.1-29.1%) | NG 76.4%, X 14.0%, W 5.9%, Y 2.0%, C 1.0%; A 0% |
| Alemayehu *et al.* (2017)(47) | Central Ethiopia (Addis Ababa, school children, n=240) | 2016 | NP | 20.4% (15.5-26.0%) | W 40.8%, C 24.5%, NG and others |
| Tefera Z. *et al.* (2020) (49) further analysed by Belachew T *et al.* (2023) (52) | Northwest Ethiopia (Gondar town, School children, n=524) | 2019 | OP | *N. meningitidis*: 10.1% (7.6-12.8%)<br>*M. catarrhalis*: 6.9%,<br>*N. lactamica*: 2.7% | A 24.5%*, Y/W 20.7%, B 7.6%, NG 47.2% |
| Assefa S. *et al.* (2022)(48) | Southwestern Ethiopia (Jimma, Prison, n=550) | 2019 | OP | 13.8% (7.20-18.20%) | NG 34.2%, W/Y 28.9% |
| Belayneh *et al.* (2026)(50) | Northwest Ethiopia (Gondar, High School & University Students, n=1,025) | 2019 | OP | 1.9% (1.1-2.9%) | NG/cnl 84.2%, W 10.5%, B 5.3%; A 0% |
| Birhanu *et al.* (2024) (53) | Northwest Ethiopia (Gondar town, Gondar Hospital Paediatric Outpatients, n=385) | 2023 | NP | *N. meningitidis*: 8.8%,<br>*M. catarrhalis*: 10.6%,<br>*H. influenzae*: 6.8% | Not serogrouped |

CI, confidence interval; NG, non-groupable; NP, nasopharyngeal; OP, oropharyngeal. \* Serogroup A was reported by seroagglutination or latex typing of uncertain reliability and was not confirmed by molecular methods

As with the surveillance data, existing carriage data are insufficient to inform current vaccine policy. Reported prevalence varies widely, highlighted by a fivefold discrepancy between two studies sampling overlapping school-age populations in the same year. This variation stems from differences in sampling sites, culture methods, and isolate identification techniques. Furthermore, the 2019 report of 24.5% MenA carriage (proportion of identified serogroups) in Gondar (49) is inconclusive; concurrent (50) and pre-vaccination studies found no MenA carriage, and no post-campaign MenA cases have been confirmed nationally. Moreover, post-campaign studies lack methodological rigour: all use convenience sampling without defined denominators, omit the general 1–29-year age group and high-risk western lowlands, and lack genomic characterisation to distinguish carriage from hypervirulent epidemic lineages. Consequently, Ethiopia lacks population-representative, molecularly confirmed, and lineage-characterised carriage data for the post-MenAfriVac era. Addressing this gap requires a multisite, population-based study covering the 1–29 age group across high and low-risk regions, applying standardised sampling, molecular confirmation, and genogrouping. Such a study would identify circulating serogroups and provide policymakers with the evidence needed to guide vaccine strategy.

### Population immunity and susceptibility

#### i. Seroepidemiology

All published serological studies from Ethiopia predated the vaccine and targeted different populations, so they cannot inform current policy on long-term population immunity or the effectiveness of the MenAfriVac programme. Existing studies investigate the immunology of natural infection (54) and baseline pre-vaccine seroprevalence (55) but none estimates current population immunity following the MenAfriVac programme. A targeted, multicentre serological study could therefore help characterise current population susceptibility across Ethiopia. Serosurveys done after MenAfriVac rollout elsewhere in the meningitis belt offer a useful preview of what an Ethiopian study could show. A serosurvey conducted in Burkina Faso one year after the 2010 campaign revealed high seroprotection among vaccinated age groups, with antibody titers significantly exceeding pre-vaccination levels (56). However, subsequent evidence showed that antibody titers wane over time, most rapidly in individuals vaccinated during early childhood, highlighting the potential need for booster campaigns (6, 57). The lesson for Ethiopia is that the vaccine produces a strong initial antibody response that then wanes unevenly by age, which is exactly the kind of data required to inform the timing of a booster in Ethiopia, where the baseline seroprevalence was the lowest among all meningitis belt countries (55).

Interpreting serological data in the meningitis belt presents methodological challenges. Historical protective thresholds, derived from European polysaccharide vaccine trials (58) and correlate of protection defined in US Army recruits by Goldschneider et al. (59), may not directly apply to serogroup A in this region, where high seroprevalence often reflects cumulative exposure rather than validated immunity (60). Consequently, results from serosurveys must be interpreted alongside carriage and disease surveillance evidence.

A further question in seroepidemiological studies is which assay to use. The serum bactericidal antibody (SBA) assay, which measures functional killing, is the gold-standard correlate of protection for meningococcal disease (61) but requires specialised laboratory capacity, including complement sourcing and standardised target strains, that is currently limited in Ethiopia. Enzyme-Linked Immunosorbent Assay (ELISA), by contrast, is feasible in low-resource settings and provides a quantitative binding measure that serves as a proxy for population-level exposure and immune status (7, 55, 56). In Ethiopia, ELISA could measure seropositivity, characterise immunity in the unvaccinated population, and build the initial evidence base for vaccine strategy, particularly given that current clinical surveillance cannot provide this information.

#### ii. MenAfriVac campaign coverage

MenAfriVac reached approximately 61 million people, 98.3% of the 62 million target population aged 1 to 29 years (43) (Table 4).

**Table 4:**
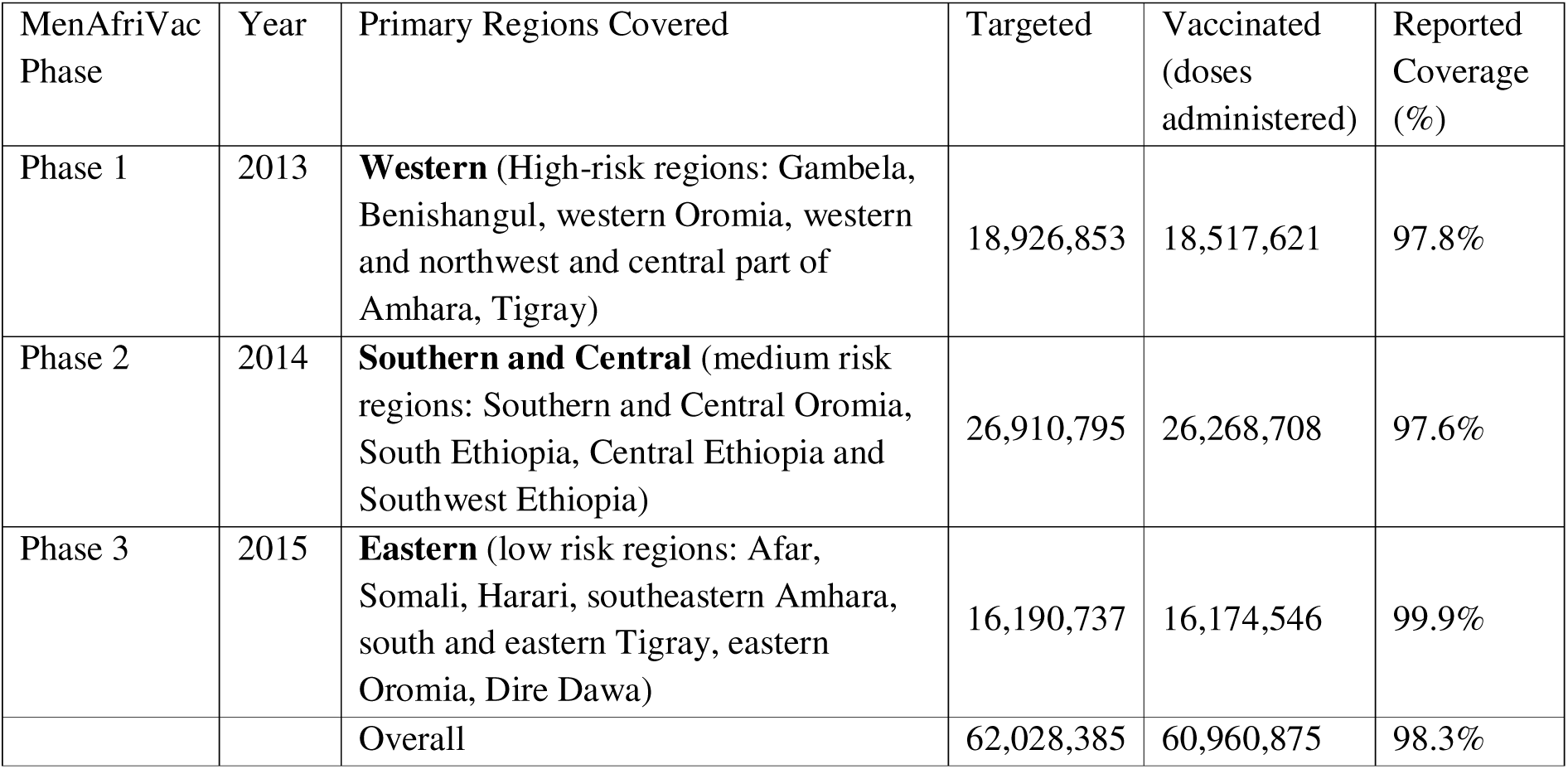
Geographical areas and population covered by three phases of the MenAfriVac campaign.

#### iii. The modelled MenA immunity gap

Waning immunity starting in 2014 among individuals vaccinated in 2013 limited overall MenA protection to an 85% peak by 2015 (Figure 6). The model shows that while cohort replacement gradually reduces protection, waning immunity within the vaccinated population is the primary driver of the overall decline in protection over time. Subsequently, the proportion of individuals protected steadily declined, reaching 41% in 2020. These projections indicate a further decline to 24% by 2024 and, without intervention, to 11% by 2030 (Figure 6).

**Figure 6:**
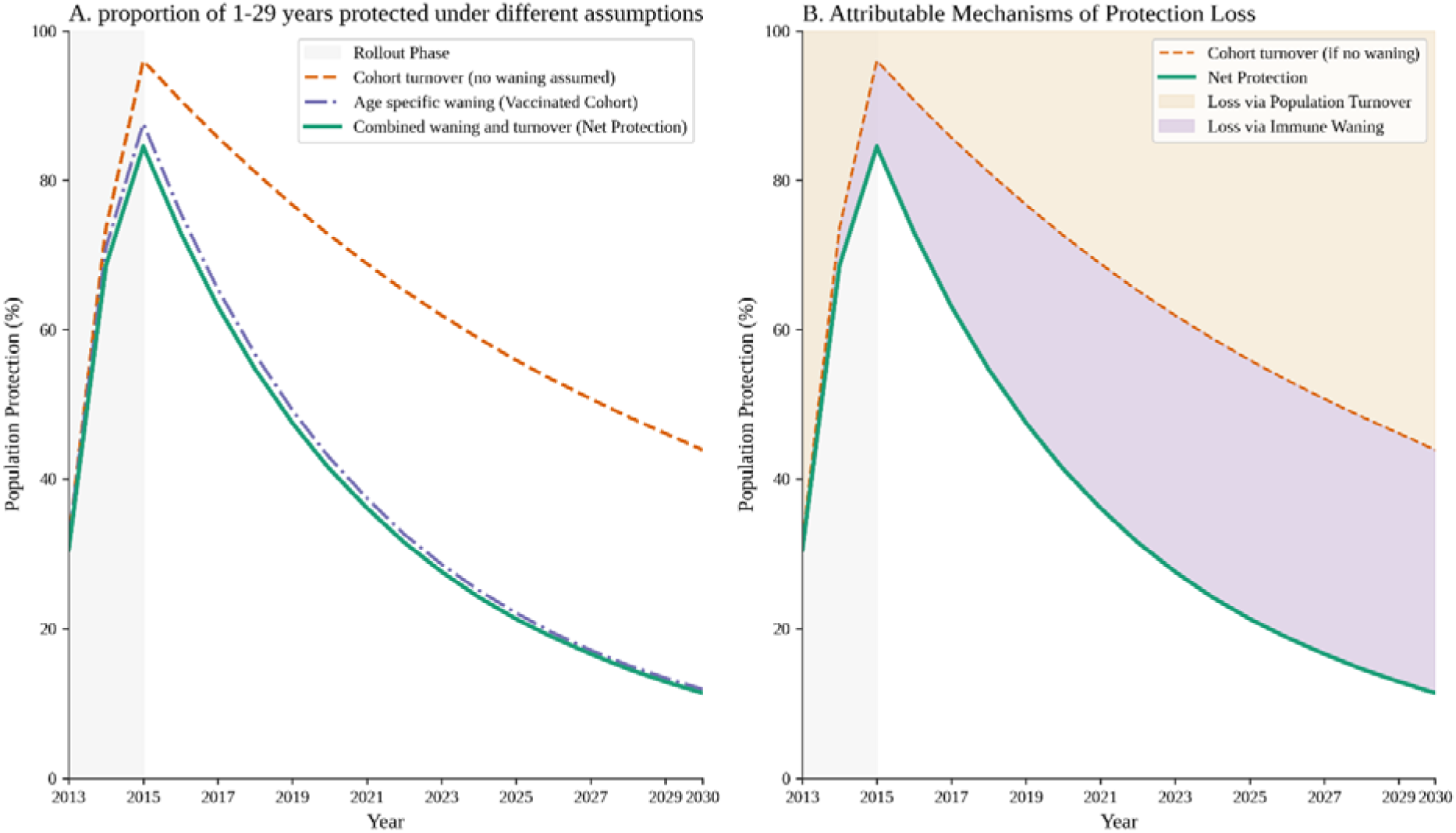
Proportion of targeted population (1-29 years) protected against MenA following mass vaccination with MenAfriVac in Ethiopia (trends from 2013 to 2025 and forward projection to 2030 in the absence of further vaccination campaigns). (A) Projected population protection (%) under individual and combined mechanisms of cohort turnover and age-specific immune waning (grey shading: 2013– 2015 rollout). (B) Relative contributions of immune waning and population turnover to total protection loss over time

Ethiopia’s population protection reached 85% nationwide in 2015, then declined to under 40% in central and western regions and below 60% in the east by 2021. By late 2025, protection dropped below 40% across nearly all regions. Without renewed vaccination efforts, protection is expected to fall below 20% by 2030, with the western region facing the highest risk (Figure 7).

**Figure 7:**
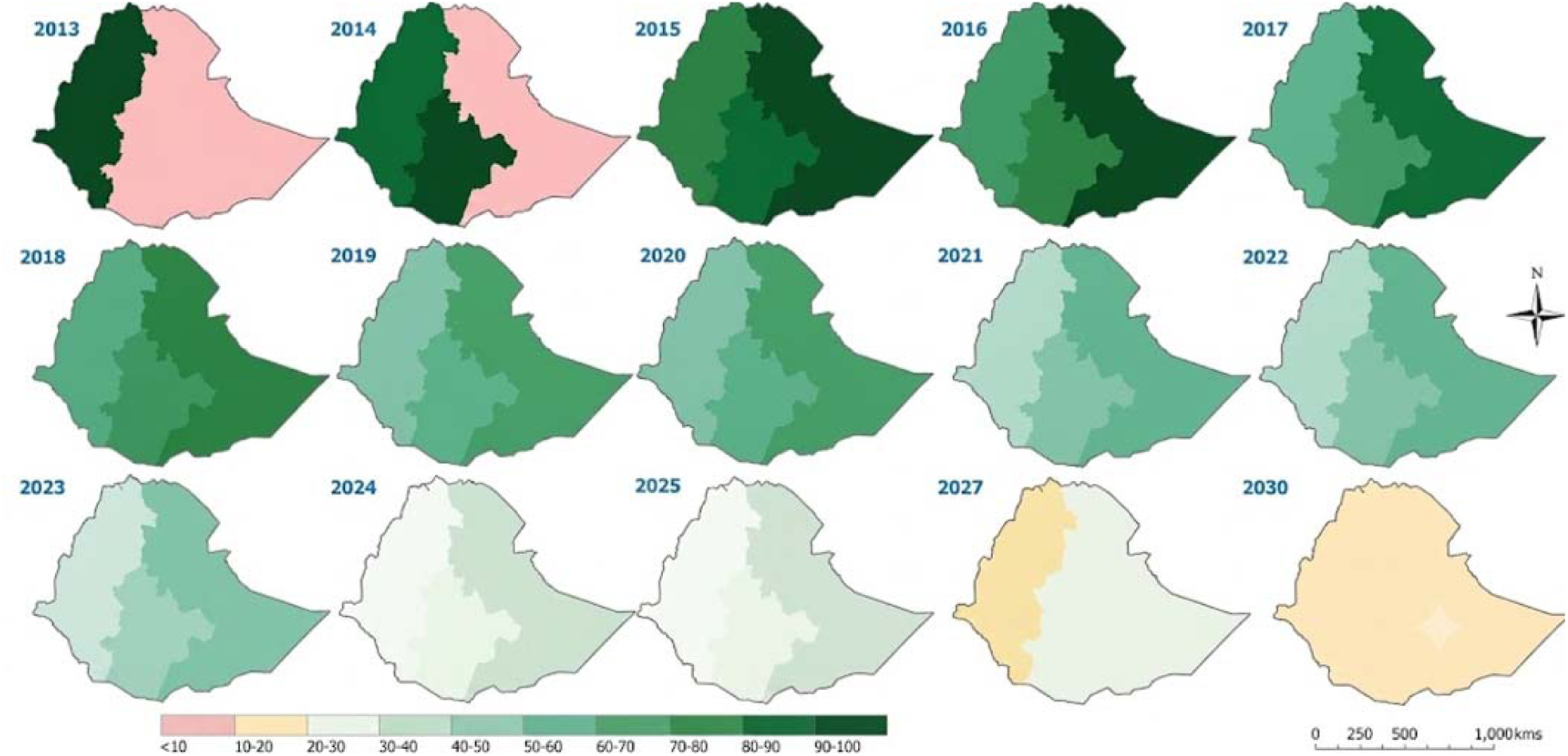
Proportion of the population protected (1-29 years) against MenA due to MenAfriVac vaccine over administrative zones of Ethiopia from 2013 to 2030.

While providing a useful risk-assessment baseline, the model may underestimate true population susceptibility because of data constraints. First, administrative coverage is frequently overstated through numerator inflation from over-reporting, double-counting or off-target vaccination (62). Second, the model lacks age-specific disaggregation, making it difficult to reconstruct cohort-specific coverage. Third, the denominators may be underestimated because projections relied on an outdated 2007 census with assumed mortality rates, which biases both coverage estimates and current population counts.

### The Policy environment

#### i. Current national strategies, policies, and guidelines

Prior to the introduction of MenAfriVac, Ethiopia managed meningococcal disease primarily through reactive vaccination using serogroup A/C polysaccharide vaccines, targeted chemoprophylaxis for close contacts, and clinical case management (34, 63). While aligned with contemporary guidelines, this strategy had clear limitations: polysaccharide vaccines provided weak, short-lived immunity and lacked the preventive efficacy of conjugate vaccines. To strengthen monitoring, the MoH joined the WHO Paediatric Bacterial Meningitis (PBM) surveillance network in 2002 (9). Following the 2013 introduction of MenAfriVac, Ethiopia transitioned from reactive epidemic control to proactive disease prevention by adopting a strategy built on mass vaccination, strengthened surveillance, and standardized outbreak management guidelines (14).

#### ii. Actors and contexts

Ethiopia’s EPI is led by the Federal Ministry of Health with technical guidance from the E-NITAG (20). International organisations including Gavi, the Vaccine Alliance, PATH and WHO supported the meningococcal A vaccine programme in Ethiopia from development through funding to implementation (64). Gavi contributed more than US$78 million to the MenAfriVac campaign (64). WHO supports vaccine programmes through safety monitoring, prequalification and policy development, and it backed the MenAfriVac campaign through the Meningitis Vaccine Project, its partnership with PATH (65). A new pentavalent meningococcal conjugate vaccine (Men5CV), protecting against serogroups A, C, W, Y and X in a single dose, was developed by PATH and the Serum Institute of India and was prequalified by WHO in July 2023. In September 2023, the SAGE recommended that Men5CV be introduced into the routine programmes of all 26 belt countries at 9-18 months of age, replacing existing MenA conjugate vaccine where used. It also urged countries that have not yet added MenA conjugate vaccine to routine immunisation to do so now rather than wait for Men5CV, given the continuing resurgence risk. WHO issued the corresponding position paper in January 2024 (21).

In its sixth investment strategy (2026–30), Gavi separated vaccine support into guaranteed budgets for existing routine programmes and discretionary budgets allocated against assessment criteria; meningococcal vaccines are funded from the discretionary budget (22). Support is tiered by national risk classification. Countries classified as high risk are supported to transition from MenAfriVac to Men5CV in routine immunisation, together with a one-off preventive campaign timed to coincide with the switch. Low– and medium-risk belt countries are supported to introduce MenAfriVac only, until their risk profile or financing position changes. Ethiopia, a Gavi-supported country since 2002 (66), therefore faces a two-step pathway: it must first introduce MenAfriVac into routine immunisation and complete a risk assessment defining its high-risk areas before it can apply for Men5CV. Current UNICEF reference prices are US$0.98 per dose for MenAfriVac and US$3.00 per dose for Men5CV (67). Should surveillance data remain scarce, Ethiopia risks classification as low-to-medium risk, which would limit it to MenAfriVac.

#### iii. Vaccine policy prioritisation and the E-NITAG

In 2021, Ethiopia outlined a strategic expansion of its EPI to include the MenAfriVac vaccine, Hepatitis B birth dose, Measles-Rubella (MR), a second dose of Inactivated Poliovirus Vaccine (IPV2), and Yellow Fever (68). The Hepatitis B birth dose and IPV2 were subsequently integrated into the routine schedule (69) while MenAfriVac was not. The published E-NITAG record does not explain why MenAfriVac’s integration was deferred. MoH & Gavi’s country appraisal, however, indicated that the vaccine was not introduced due to insufficient local evidence and competing priorities, and noted that an ongoing risk assessment is intended to facilitate its introduction during the next strategic period (70).

A new vaccine introduction is a multi-step process involving prioritisation, planning, and implementation. This process is guided by E-NITAG. Previously, E-NITAG reviewed each new vaccine introduction individually, resulting in a recommended list of several new vaccines to be introduced over a short period without conducting a preliminary prioritisation exercise. This approach has sometimes led to delays in vaccine introduction even when an E-NITAG recommendation had already been issued. Since 2024, E-NITAG has applied the Multi-Criteria Decision Analysis (MCDA) approach through the New Vaccine Introduction Prioritization and Sequencing Tool (NVI-PST) (20). In March 2025, E-NITAG scored six shortlisted candidate vaccines across importance and feasibility criteria. The multivalent meningococcal conjugate vaccine scored high on public health importance, ranking first in mortality, second in incidence, and third overall. However, a poor feasibility score relegated it to the medium-priority group as important but less feasible (Table 5).

**Table 5:** E-NITAG Prioritisation Results for the 2026–2030 National Immunisation Strategy.

| Vaccine candidate | Ranking: importance/ feasibility /combined | Priority | Sequencing | Stated rationale (20) |
| --- | --- | --- | --- | --- |
| Hexavalent (DTwP-HepB-Hib plus IPV) | 1.9 / 2.4 / 2.1 | High | 2026–2027 | Modestly higher procurement cost offset by less cold chain space, fewer injections and reduced health-worker time. |
| Rubella (as measles-rubella) | 2.8 / 2.9 / 2.8 | High | 2026–2027 | No additional cold chain space or workforce time relative to the existing measles vaccine; disease burden estimated from modelled congenital rubella syndrome data in the absence of reliable national estimates. |
| Multivalent meningococcal (Men5CV) | 2.8 / 4.3 / 3.3 | Medium | 2028–2030 | Important but less feasible. Ranked first of six on mortality and lethality and second on incidence. MenAfriVac is a prerequisite for Gavi funding eligibility for transition. |
| RSV (maternal) | 3.7 / 4.9 / 4.1 | Medium | 2028–2030 | Ranked least feasible of the six. Not included under Gavi co-financing |
| Cholera (OCV) | 4.7 / 3.6 / 4.3 | Low | Not sequenced | Recommendation covers vaccines for routine use. |
| Typhoid (TCV) | 5.1 / 2.9 / 4.4 | Low | Not sequenced | Ranked lowest of the six on importance, despite comparatively favourable feasibility. |

Several features of the E-NITAG scoring framework are likely to have systematically disadvantaged epidemic-prone pathogens, including meningococcus. Critically, MenAfriVac was excluded from the shortlisted candidates, leaving the exercise without a recommendation for its introduction. The framework’s underlying criteria further disadvantaged the vaccine by evaluating disease importance strictly through annual incidence, prevalence, and severity, while ignoring epidemic potential and the value of averting low-frequency, high-consequence outbreaks. Under this design, a pathogen causing 45,000–50,000 cases in a single epidemic but near-zero cases between outbreaks would be misclassified as low-burden (1, 33, 38). Additionally, the framework omitted protection duration, masking critical gaps in population-level immunity. Although serogroup coverage was recognised as a crucial parameter, it was also left unranked because non-meningococcal candidates rendered it non-discriminatory; while methodologically defensible, this decision eliminated the sole criterion where a meningococcal vaccine held a clear comparative advantage.

Beyond disease burden, the framework’s handling of economic feasibility introduced a further blind spot. Men5CV was scored on programme costs, unit prices and Gavi co-financing as a standalone candidate, with no intra-class comparison against MenAfriVac. The two are not competing options but sequential steps: routine MenAfriVac introduction is a precondition for Gavi eligibility for Men5CV, and MenAfriVac costs roughly a third of Men5CV’s per-dose price (67). This comparison helps Ethiopia decide whether to introduce the lower-cost prerequisite product into routine immunisation before a resurgence occurs.

These design features were compounded by data limitations. E-NITAG cited severe gaps in data availability and quality at national and regional levels and called for centralised repository access within surveillance systems. The prioritisation entirely relied on global modelled estimates that are themselves driven by unrepresentative local surveillance data, a circular dependency E-NITAG acknowledged but could not resolve within the scope of the assessment.

## Implications

Transitioning from reactive outbreak response to routine integration within the EPI offers Ethiopia a sustainable route to maintaining population immunity and forestalling epidemic resurgence. This situational analysis, however, identifies two bottlenecks to that transition: a surveillance system that cannot generate the evidence a vaccine decision requires, and a prioritisation framework that is structurally insensitive to the form of risk meningococcal disease presents.

First, national meningitis surveillance is geographically and diagnostically inadequate for guiding vaccine policy. Geographically, the three sentinel sites are urban and located in central and north-western Ethiopia. None of these covers the western epidemic regions or the eastern pastoralist lowlands. Diagnostically, confirmation fails at every step of the clinical pathway: blood and CSF cultures are rarely performed and serogrouping is almost absent after 2017. Because notification depends entirely on unconfirmed clinical diagnosis, the system can neither quantify true burden nor detect serogroup replacement. The apparent absence of MenA since 2017 is therefore uninterpretable.

Second, the E-NITAG prioritisation framework contains internal logic gaps that risk delaying vaccine introduction. The framework rated Men5CV a medium priority for 2028–2030 but omitted MenAfriVac from the candidate list, even though routine MenAfriVac introduction is a prerequisite for Gavi co-funding of Men5CV. Prioritising Men5CV without an operational schedule for routine MenAfriVac introduction therefore creates a sequencing bottleneck that makes the 2028–2030 target unachievable. The framework also scores disease importance on annual incidence, prevalence and severity alone. A pathogen capable of 45,000–50,000 cases in a single epidemic (1, 33, 38) but with near-zero inter-epidemic incidence is consequently misclassified as low burden, and duration of protection, the parameter that determines when a vaccinated population becomes susceptible again, is not scored at all.

Three actions follow, with two distinct purposes: to rebuild the evidence base that meningococcal vaccine policy in Ethiopia currently lacks, and to act on the resurgence risk. First, Ethiopia should redesign meningitis surveillance as a nested three-tier system adapted from existing vaccine-preventable disease platforms. At Tier 1, all primary hospitals and primary health care units (PHCUs) would report suspected cases through DHIS2, establishing population-level clinical risk. At Tier 2, a defined subset of regional and general hospitals would collect CSF and perform culture and PCR to quantify confirmed bacterial meningitis. At Tier 3, a geographically representative set of referral hospitals, deliberately including sites in the western high-risk zones and the eastern lowlands, both currently unrepresented, would perform molecular serogrouping and genomic characterisation, supported by the national reference laboratory network. Each tier draws from the catchment of the one below it, so that clinical, aetiological and serogroup data describe the same population rather than three unrelated ones.

Second, decision-makers should establish a pathway, separate from the current prioritisation cycle, to reintroduce MenAfriVac into routine EPI with an accompanying catch-up campaign. This is consistent with Gavi’s country appraisal recommendation and current SAGE guidance, addresses a projected fall in population protection to 11% by 2030, and simultaneously satisfies the eligibility prerequisite for Men5CV. Third, because a redesigned surveillance system will take several years to yield policy-grade data, interim multisite carriage and seroepidemiological studies are required to characterise circulating serogroups and residual population immunity in time for the next strategy review. E-NITAG should use that review to amend its scoring criteria so that duration of protection and epidemic potential are explicitly weighted, preventing the recurrent misclassification of epidemic-prone pathogens.

## Conclusion

In summary, Ethiopia cannot determine whether MenA protection has waned to levels that would permit epidemics, or whether non-A serogroups are circulating, because its surveillance system is not built to answer either question. Strengthening the meningococcal evidence is the precondition for every subsequent vaccine decision. It requires a nested three-tier system in which primary facilities report clinical cases, regional and general hospitals confirm aetiology by culture and PCR, and a geographically representative set of referral hospitals performs serogrouping and genomic characterisation. As such a system will take several years to yield policy-grade data, periodic carriage and seroepidemiological studies remain necessary. Meanwhile, with modelled protection projected to fall over time, MenAfriVac should be reintroduced into routine immunisation without waiting for further prioritisation activities.

## Acknowledgments

We thank the Ministry of Health for providing the vaccine uptake data and the Ethiopian Public Health Institute (EPHI) for providing the disease surveillance data.

## Data availability statement

The datasets generated and/or analysed during the current study, along with the code used for analysis, are available from the corresponding author upon reasonable request.

